# Real-world data suggest antibody positivity to SARS-CoV-2 is associated with a decreased risk of future infection

**DOI:** 10.1101/2020.12.18.20248336

**Authors:** Raymond A. Harvey, Jeremy A. Rassen, Carly A. Kabelac, Wendy Turenne, Sandy Leonard, Reyna Klesh, William A. Meyer, Harvey W. Kaufman, Steve Anderson, Oren Cohen, Valentina I. Petkov, Kathy A. Cronin, Alison L. Van Dyke, Douglas R. Lowy, Norman E. Sharpless, Lynne T. Penberthy

**Author notes:** **Corresponding Author:** Lynne T. Penberthy, MD, MPH; National Cancer Institute, National Institutes of Health, 9609 Medical Center Dr, Rockville, MD 20850.

## Abstract

**Importance:** There is limited evidence regarding whether the presence of serum antibodies to SARS-CoV-2 is associated with a decreased risk of future infection. Understanding susceptibility to infection and the role of immune memory is important for identifying at-risk populations and could have implications for vaccine deployment.

**Objective:** The purpose of this study was to evaluate subsequent evidence of SARS-CoV-2 infection based on diagnostic nucleic acid amplification test (NAAT) among individuals who are antibody-positive compared with those who are antibody-negative, using real-world data.

**Design:** This was an observational descriptive cohort study.

**Participants:** The study utilized a national sample to create cohorts from a de-identified dataset composed of commercial laboratory test results, open and closed medical and pharmacy claims, electronic health records, hospital billing (chargemaster) data, and payer enrollment files from the United States. Patients were indexed as antibody-positive or antibody-negative according to their first SARS-CoV-2 antibody test recorded in the database. Patients with more than 1 antibody test on the index date where results were discordant were excluded.

**Main Outcomes/Measures:** Primary endpoints were index antibody test results and post-index diagnostic NAAT results, with infection defined as a positive diagnostic test post-index, as measured in 30-day intervals (0-30, 31-60, 61-90, >90 days). Additional measures included demographic, geographic, and clinical characteristics at the time of the index antibody test, such as recorded signs and symptoms or prior evidence of COVID-19 (diagnoses or NAAT+) and recorded comorbidities.

**Results:** We included 3,257,478 unique patients with an index antibody test. Of these, 2,876,773 (88.3%) had a negative index antibody result, 378,606 (11.6%) had a positive index antibody result, and 2,099 (0.1%) had an inconclusive index antibody result. Patients with a negative antibody test were somewhat older at index than those with a positive result (mean of 48 versus 44 years). A fraction (18.4%) of individuals who were initially seropositive converted to seronegative over the follow up period. During the follow-up periods, the ratio (CI) of positive NAAT results among individuals who had a positive antibody test at index versus those with a negative antibody test at index was 2.85 (2.73 - 2.97) at 0-30 days, 0.67 (0.6 - 0.74) at 31-60 days, 0.29 (0.24 - 0.35) at 61-90 days), and 0.10 (0.05 - 0.19) at >90 days.

**Conclusions:** Patients who display positive antibody tests are initially more likely to have a positive NAAT, consistent with prolonged RNA shedding, but over time become markedly less likely to have a positive NAAT. This result suggests seropositivity using commercially available assays is associated with protection from infection. The duration of protection is unknown and may wane over time; this parameter will need to be addressed in a study with extended duration of follow up.

**Key Points:** *Question:* Can real-world data be used to evaluate the comparative risk of SARS-CoV-2 infection for individuals who are antibody-positive versus antibody-negative?

*Finding:* Of patients indexed on a positive antibody test, 10 of 3,226 with a NAAT (0.3%) had evidence of a positive NAAT > 90 days after index, compared with 491 of 16,157 (3.0%) indexed on a negative antibody test.

*Meaning:* Individuals who are seropositive for SARS-CoV-2 based on commercial assays may be at decreased future risk of SARS-CoV-2 infection.

## Introduction

Since the emergence of SARS-CoV-2 in late 2019, limited research has shown that the majority of patients who clear their infections develop serum antibodies against the virus that last for at least several months^1–6^ but may decline over time.^7^ Although it has been speculated that the development of antibodies may be associated with a decreased risk of reinfection, the evidence for this hypothesis is limited and often anecdotal.^8,9^ Furthermore, documented reports of reinfection in patients with SARS-CoV-2 antibodies have raised the possibility that seropositivity might be associated with limited protection against different viral strains.^10–14^ SARS-CoV-2 infected individuals may also shed viral RNA without producing live virus for 12 weeks or more after resolution of symptoms,^15–20^ making it challenging to distinguish reinfection from prolonged RNA shedding. As the pandemic continues, understanding the role of serostatus on the potential for infection is critical, as it may drive choices of personal behavior and expectations around herd immunity. It might also help inform the challenging policy decisions surrounding the prioritization of vaccine supplies.

Commercially available antibody assays, with their high sensitivity and low false-positive rate,^21–23^ serve as a useful marker of prior SARS-CoV-2 infection, but to date, their ability to predict risk of future infection is unknown. Given the critical lack of data in this area, the CDC currently recommends that individual serology results not be used for any decision-making regarding personal behavior (such as return to work decisions, use of PPE, and social distancing). These gaps highlight the clear need for generalizable data that can assess the impact of seropositivity on risk of future infection. Real world data (RWD) that is available longitudinally at the individual level offers the possibility to study the experiences of a seropositive COVID-19 population in near-real time, while maximizing sample size and observability over time.

In this paper, we employ a RWD approach to investigate the relationship between SARS-CoV-2 antibody status and subsequent NAAT results, in an effort to understand how serostatus may predict risk of reinfection.

## Methods

In this retrospective observational descriptive cohort study, we used de-identified individual-level laboratory testing data provided by HealthVerity (Philadelphia, PA), a for-profit data aggregator providing access to linked data from 70 different commercial health data sources. Data available for this study included clinical commercial laboratory results from several national and regional laboratories, representing more than 50% of commercial antibody and diagnostic testing in the United States. These longitudinally linked commercial lab data were the primary data sources for the analyses in this study. In addition, longitudinal data were captured on each individual from open and closed medical and pharmacy claims, electronic health record data, and hospital billing data from multiple vendors (see details in Supplementary Materials). These data served to assess the availability of data to characterize patient level comorbid conditions and other risk factors that might impact risk of infection and outcome. The data derived from laboratories and medical record systems as well as insurance claims cover the US but may under-sample the Midwest region. To create the consolidated, de-identified dataset with longitudinal patient views, all data partners used the HealthVerity technology within their system to create a unique, secure, encrypted, and non-identifiable patient token from identifiable information. This token was then employed as a consistent linkage key across datasets, and allowed for follow-up of patients who, for example, used multiple laboratory providers. No PHI (protected health information) or PII (personal identifying information) left the data owner’s possession, and all research data were certified HIPAA compliant by expert determination.

The commercial laboratories antibody testing includes a limited set of high throughput antibody tests with validation against a known standard providing between 98% to 100% agreement with both known antibody-positive and antibody-negative specimens, with a 95% confidence interval of 99-100% agreement. An evaluation of the FDA emergency use authorization (EUA) documents shows that the composite negative validation data demonstrate a 95% confidence interval range of 99% to 100%.^22,23^ Tests performed in these commercial laboratories represent tests specific for IgG, IgA, or IgM as well as those that detect multiple immunoglobulin types, although the majority of tests performed during the study period were IgG (>91%).

We examined records from December 1, 2018 through August 26, 2020 and identified individuals with a recorded SARS-CoV-2 antibody test on or after January 2020. Patients entered the cohort on the day of their first recorded antibody test, which was defined as the index date (see Figure 1). Individuals who had more than 1 antibody test with discordant results on the index day were excluded. Using our linked longitudinal dataset, we assessed demographic and geographic characteristics at index, as well as evidence of prior SARS-CoV-2 infection and key associated clinical characteristics and comorbidities. These characteristics were measured as recorded in the EHR, administrative claims, and hospital records.

**Figure 1.**
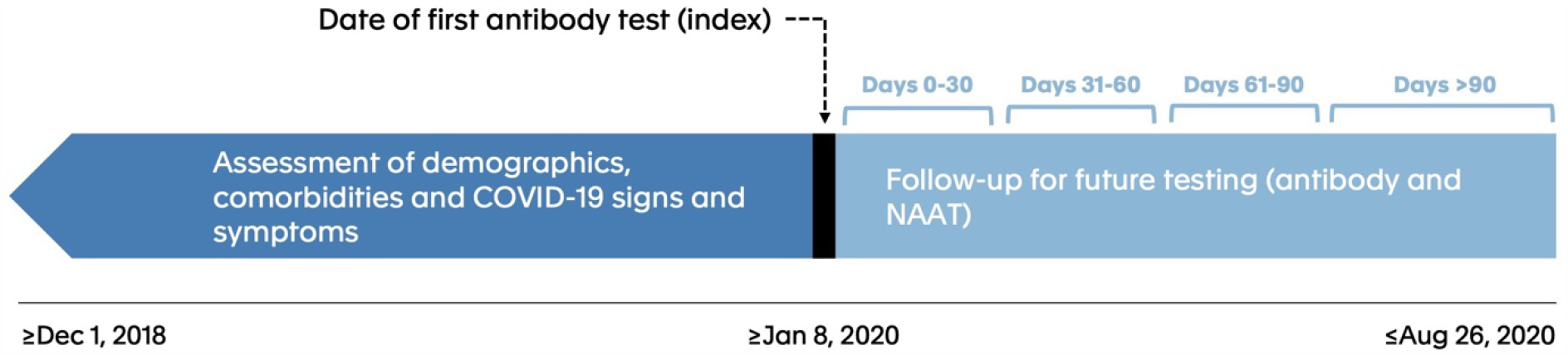
Diagram of study design. This figure shows the key elements of the study design. The study index date for each patient was the day of the patient’s first observed SARS-CoV-2 antibody test on or after January 8, 2020. Follow-up occurred in 30-day increments after the index date.

We characterized patients’ initial antibody test results as positive, negative, or inconclusive, and created 3 associated groups. We then followed patients to the end of available data (August 26, 2020) to identify further antibody testing and/or nucleic acid amplification test (NAAT) diagnostic testing, looking in 30-day intervals (0-30, 31-60, 61-90, >90 days). Within each interval and for each of the 3 index antibody groups, we assessed both the frequency of subsequent antibody or NAAT diagnostic testing and the test results. An individual was characterized as positive for an antibody or NAAT during a time period if they had at least 1 positive test during that period. Patients were counted uniquely within each time period and could have been included in multiple time periods. All analyses were done on the Aetion Evidence Platform (Aetion, Inc., New York, NY), version R4.11.

## Results

A total of 3,257,478 unique patients with an index antibody test were identified after excluding 132 patients with discordant antibody tests on the index day. Of these, 2,876,773 (88.3%) had a negative index antibody result (seronegatives), 378,606 (11.6%) had a positive index antibody result (seropositives), and 2,099 (0.1%) had an inconclusive index antibody result (sero-uncertain). (Table 1). As the sero-uncertain group was a small fraction of the study population, further reported results focus only on the seropositive and seronegative groups. Approximately 55% in each group were female. The index seronegative group was somewhat older than the index seropositive group (mean of 48 versus 44 years). A higher proportion of index seropositives resided in the Northeast United States, with fewer in the rest of the country (Figure S1 Supplemental Materials).

**Table 1.** Baseline and Pre-Index Characteristics.

| Index (First) Antibody Test Result<br>Total (n=3,257,478) |  |  |  |
| --- | --- | --- | --- |
|  | Negative Result<br>2,876,773 (88.3%) | Positive Result<br>378,606 (11.6%) | Inconclusive Result<br>2,099 (0.1%) |
| **Demographic Characteristics** |  |  |  |
| **Age** |  |  |  |
| Mean (SD) | 47.66 (17.63) | 44.34 (18.09) | 49.45 (19.22) |
| Median [IQR] | 48.00 [34.00, 61.00] | 45.00 [30.00, 58.00] | 50.00 [35.00, 64.00] |
| **Sex** |  |  |  |
| Female, n (%) | 1,599,898 (56.7%) | 202,157 (54.1%) | 1,143 (55.3%) |
| **Geographic Region** |  |  |  |
| Northeast, n (%) | 1,008,720 (35.8%) | 230,513 (61.7%) | 305 (14.8%) |
| Midwest, n (%) | 239,837 (8.5%) | 16,735 (4.5%) | 56 (2.7%) |
| South, n (%) | 786,551 (27.9%) | 51,648 (13.8%) | 403 (19.5%) |
| West, n (%) | 514,441 (18.2%) | 26,706 (7.1%) | 1,066 (51.6%) |
| **Index Antibody Test Type** |  |  |  |
| Antibody; n (%) | 237,035 (8.2%) | 49,414 (13.1%) | 3 (0.1%) |
| Antibody IgA; n (%) | 1,782 (0.1%) | 50 (0.0%) | 19 (0.9%) |
| Antibody IgG; n (%) | 2,625,428 (91.3%) | 328,506 (86.8%) | 1,648 (78.5%) |
| Antibody IgM; n (%) | 12,528 (0.4%) | 636 (0.2%) | 429 (20.4%) |
| **Patient Comorbidities** |  |  |  |
| **Chronic Conditions\*** |  |  |  |
| Hypertension, n (%) | 430,516 (24.2%) | 52,700 (24.7%) | 429 (30.8%) |
| Ischemic Heart Disease, n (%) | 96,920 (5.4%) | 10,423 (4.9%) | 137 (9.8%) |
| Coronary Heart Disease, n (%) | 80,730 (4.5%) | 8,333 (3.9%) | 118 (8.5%) |
| Metabolic Syndrome, n (%) | 42,549 (2.4%) | 6,244 (2.9%) | 41 (2.9%) |
| Vitamin D Deficiency, n (%) | 219,142 (12.3%) | 30,930 (14.5%) | 145 (10.4%) |
| Overweight, n (%)^ | 267,601 (14.5%) | 32,452 (14.7%) | 223 (15.3%) |
| Obese, n (%)^ | 311,393 (16.8%) | 42,890 (19.5%) | 301 (20.7%) |
| **Pre-index COVID-19 Diagnosis\*** |  |  |  |
| Patients with Pre-index Diagnosis, n (%) | 11,305 (0.4%) | 23,824 (6.8%) | 52 (2.6%) |
| Median days [IQR] to Most Recent Pre-index Diagnosis | 1 [1, 19] | 18 [2, 38] | 10 [1, 24] |

\*claims and chargemaster
^claims, chargemaster and EHR

The seropositive and seronegative groups each had a median of 396 days of observable person-time prior to the index date. As assessed over that time, most COVID-19 signs and symptoms were similar between the seropositive and seronegative groups, although the seropositive groups had higher proportions of recorded fever (6.3% among seropositives versus 3.5% among seronegatives), acute respiratory failure (1.2% versus 0.4%), and viral infection (4.3% versus 2.0%). Other comorbidities, including were largely comparable between the seropositive and seronegative groups, with the exceptions of obesity (19.5% vs. 16.8%) and Vitamin D deficiency (14.5% vs. 12.3%) which were slightly higher among seropositives than seronegatives (Table 1). As expected, evidence of prior COVID-19 diagnosis varied across the 3 groups. Evidence of prior disease based on laboratory, claims, and/or chargemaster diagnostic codes, was 0.7% for the seronegatives, 18.4% for the seropositives, and 6.7% for the sero-uncertain group. These results indicate that seropositive individuals were more likely to have had symptoms of and/or a diagnosis of COVID than seronegative individuals, although the majority of subjects in both groups did not have evidence of prior infection in the observable data.

The linked data permitted individual longitudinal follow-up for a median of 47 days for the seronegative group (interquartile range: 8 to 88 days) and a median of 54 days for the seropositive group (IQR: 17 to 92 days). Over the available follow-up time, we examined the duration of seropositivity in the index positive cohort. Among the 378,606 patients with a positive antibody test at index, 9,895 (2.6%) had at least one subsequent antibody test during follow-up. For the index seropositive patients who were retested, 12.4% tested negative when retested within 0-30 days, increasing to 18.4% testing seronegative when the subsequent antibody test occurred >90 days after the index antibody test (Figure 2). These findings are consistent with prior studies suggesting that antibody levels wane in a modest fraction of individuals over a period of months after initial detection.^1–3^

**Figure 2.**
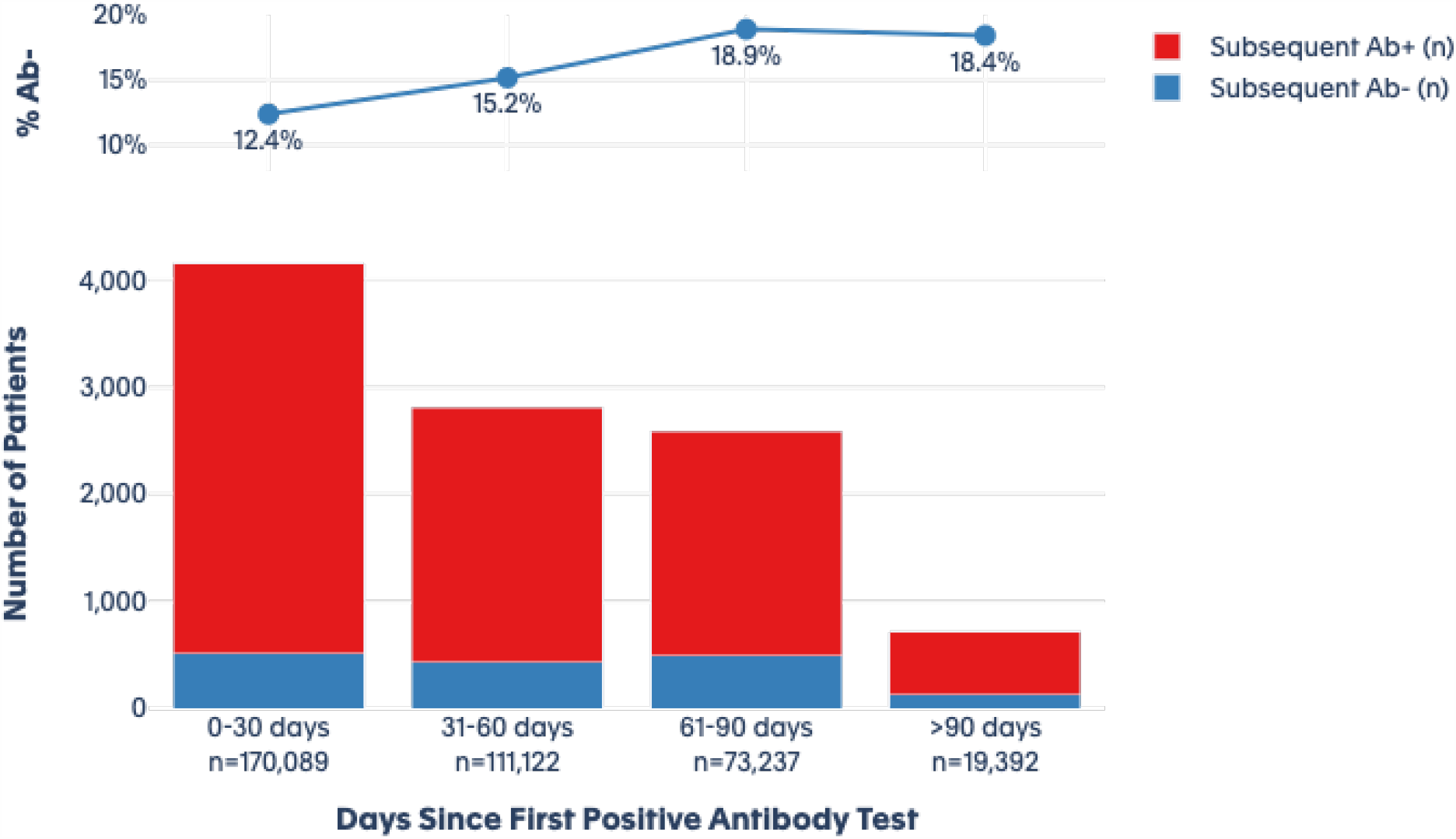
Subsequent antibody testing among index antibody positive patients over time (n = 378,606) This figure shows the results of subsequent antibody tests among the group of patients with an initial positive antibody test. Over the four time periods, blue bars represent those who subsequently test negative for antibodies, while red bars show those who subsequently test positive. The blue line shows the percentage of patients who subsequently tested negative in each time period.

We next considered the relationship between index serostatus and future NAAT testing patterns. Among the seropositive patients, 11.0% (n=41,587) had ≥1NAAT during follow-up, while among seronegatives, 9.5% (n=273,735) did so. Patients may have had multiple NAAT tests during follow-up; seropositives had a mean of 3.3 NAAT over the follow-up period, while seronegatives had 2.3 tests on average. Sero-uncertain patients were tested less frequently, with 1.5 tests per patient performed on average.

Among patients with a positive index antibody result, 3,226 (11.3%) had a positive diagnostic NAAT during follow-up that occurred within 30 days of index, decreasing consistently to 2.7% from 31-60 days, 1.1% from 61-90 days, and 0.3% at >90 days (Figure 3). For the seronegative patients, 5,638 (3.9%) showed a positive NAAT result within 30 days. That proportion remained relatively consistent at ∼3.0% over all subsequent periods of observation, including at >90 days (Figure 2). The ratio of positive NAAT results among patients who had a positive antibody test at index versus those with a negative antibody test at index declined from 2.85 [CI, 2.73 - 2.97] at 0-30 days; to 0.67 [CI 0.6 - 0.74] at 31-60 days; to 0.29 [0.24 - 0.35] at 60-90 days; and to 0.10 [CI 0.05 - 0.19] at >90 days.

**Figure 3.**
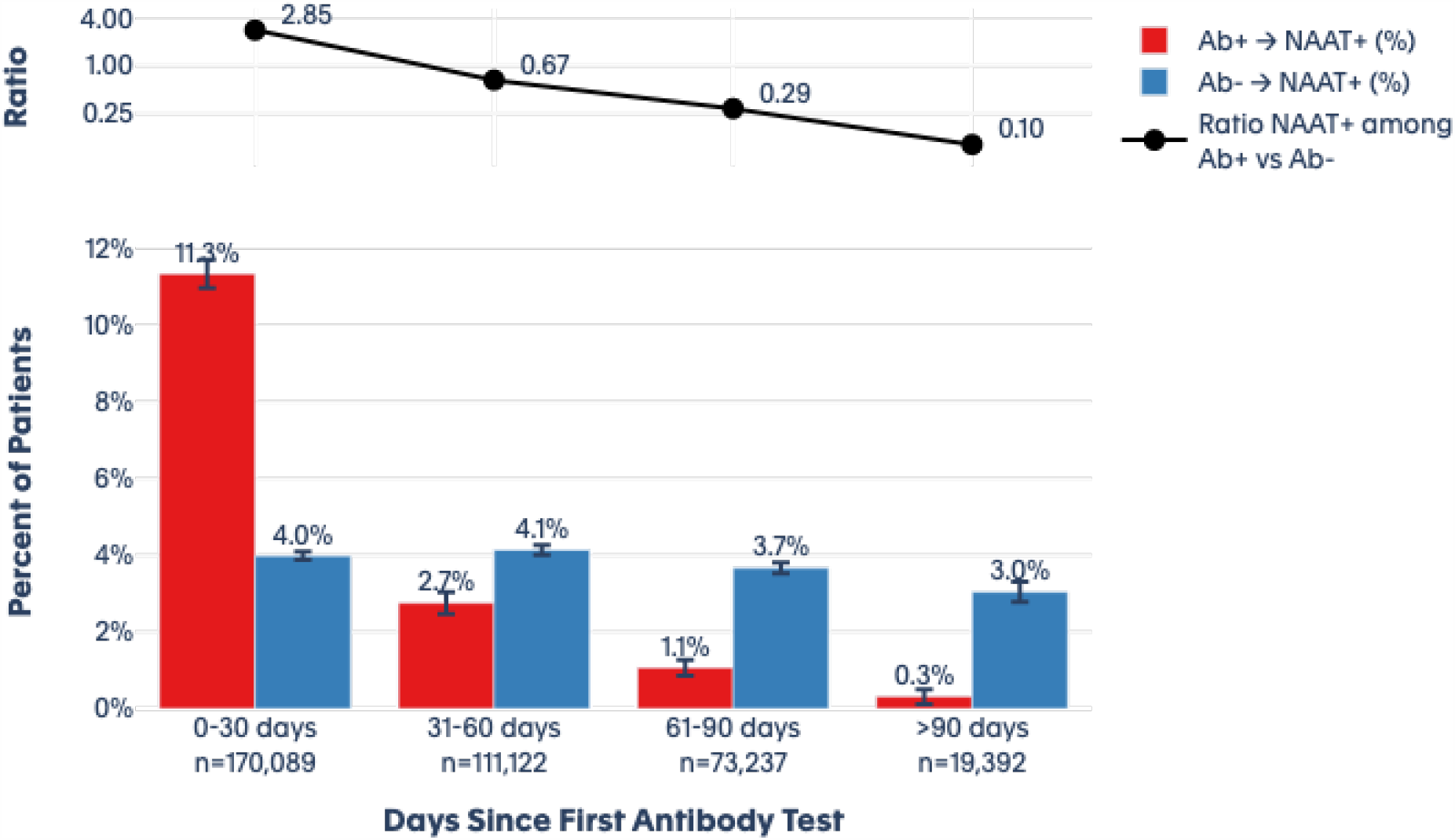
Subsequent Diagnostic (NAAT) Test Results at 30 Day Intervals. This figure shows the results of diagnostic (NAAT) testing after initial antibody testing. Over each time period, the red bars show those who test positive for the diagnostic test among those who initially tested positive for antibodies. The blue bars show those who test positive for the diagnostic test among those who initially tested negative for antibodies. The black line shows the ratio of positive diagnostic tests among those who initially tested positive for antibodies versus those who initially tested negative.

## Discussion

In this non-randomized study, de-identified RWD suggest that the presence of antibodies to SARS-CoV-2 is associated with a reduced risk of developing a subsequent positive NAAT, which may be a proxy representing a new infection or may represent continued viral shedding depending on the context and timing. While this effect was not seen in the first 30 days after an initial antibody test, the effect became pronounced after 30 days and progressively strengthened through our 90+ day observation period.

Early in the observation period, particularly in the first 30 days, positive NAAT results among seropositive patients are likely attributable to prolonged shedding of viral RNA, which should decrease through the following weeks. The increased rate of NAAT positivity observed within the first 30 days of a positive antibody test is consistent with persistent shedding of viral RNA.^15–20^ At >90 days, the vast majority of viral shedding is expected to have ceased, so positive NAAT results seen at a later interval from the index antibody test may represent new infections. False-positives are expected to be rare given the high specificity of NAAT, and are thought to generally reflect technical errors or reagent contamination (the latter is less likely due to internal controls).^24–26^

Under the assumptions that positive diagnostic tests among seronegative patients represent infections, and that positive diagnostic tests among patients who first tested seropositive >90 days prior also represent infections, we observed two notable results. First, the relatively steady ∼3.0% proportion of positive NAAT tests among index seronegative patients suggests a stable background rate of infection over the period of the study. Second, while our study was not appropriate for estimating a relative risk, the ratio of positive NAAT results among index seropositive compared with index seronegative was substantially lower — a ∼10-fold decrease — suggesting a protective effect of antibodies. While some patients may have ongoing viral RNA shedding for weeks post-infection, the sharp decline in NAAT positive test results over time in the antibody-positive cohort versus antibody-negative cohort suggests seropositive individuals are at decreased risk for future SARS-CoV-2 infection. As the pandemic infection rates varied both over time and by geographic area, we performed a preliminary stratified analysis that evaluated the risk of subsequent infection by geographic region in the United States. Although the numbers were small for some regions, the results showed a consistent decline in the ratio of NAAT positivity among seropositive versus seronegative patients in all regions over the 4 study intervals, similar to the overall analysis. This consistency supports the same level of reduction in future risk and is unlikely due to pandemic patterns of testing and/or spread (data shown in Supplemental Table S1).

The degree of protection (10-fold) associated with seropositivity appears to be comparable to that reported for the initial reports of the efficacy of mRNA vaccines in large clinical trials.^27–29^ Of course, protection induced by a safe vaccine is clearly to be preferred, as the population-wide risk of a serious outcome from an authorized or approved vaccine should be orders of magnitude lower than from natural infection.

Given the observational nature of the study, it is possible that antibody test results affected individual behavior, potentially confounding the results. We do not, however, think behavior differences likely explain the observed protection. For example, if individuals with evidence of prior infection (seropositives) were more likely to believe they possessed immunity to SARS-CoV-2, then they would be expected to engage in social behavior that placed them at <u>greater</u>, not less, risk for infection. Likewise, it is possible that seropositive individuals might be less likely to seek evaluation for subsequent symptoms of COVID, but in fact, we observed that antibody positive individuals were <u>more</u> likely to have follow-up NAAT than antibody negative individuals (3.3 vs. 2.3 subsequent tests).

We do not have insight into the clinical characteristics of the seropositive individuals who appeared to develop new infections after the index timepoint, nor could we specifically assess the clinical course of these possible infections compared with infections among the seronegative group in this study. However, some of the individuals who had NAAT positive results >60 days after an index seropositive test may represent true infections, as reinfection has been described in a small number of cases.^8–11^ Therefore on a population-wide basis, protection against reinfection is likely relative rather than complete. Factors that influence risk of reinfection — such as varying viral strains, patients’ immune status, or other patient level characteristics — should be evaluated in subsequent studies that include follow-up extended beyond the current 90 days.

There is limited but consistent evidence from two SARS-CoV-2 outbreaks suggesting that seropositivity is associated with protection from infection. In an outbreak on a fishing vessel, an attack rate of 85% was observed among the 122 individuals. Only three individuals aboard were known to have serum neutralizing antibodies prior to the outbreak, and none of them became infected.^9^ In another outbreak, at a children’s summer camp, 116/156 (76%) campers, counselors, and staff became infected, but all 24 of the individuals who were seropositive when the camp began tested negative for infection soon after the epidemic had subsided.^8^ The current findings extend those anecdotal series onto a much larger sample size based on commercially available assays used in real-world settings.

While there are acknowledged limitations to RWD, it does provide a means to complement and supplement data from clinical trials in order to formulate hypotheses and provide information on patients or clinical scenario that are not well-represented in clinical trials.^30–33^ It is particularly well-suited to situations such as an emerging pandemic, where urgent questions require rapid, near real-time answers. To be clear, however, this RWD analysis has significant limitations compared to a classical prospective seroprotection trial. It is not known whether the rate of SARS-CoV-2 exposure or pattern of longitudinal follow-up was comparable between the two groups. It is also not known if the positive NAAT results in either group were associated with clinical signs of infection. Perhaps most importantly, it is not known how long any protective effect of serostatus may last beyond the studied 90 days. These questions remain to be addressed by further research. That research can also shed light on whether a seropositive individual who subsequently becomes seronegative may be associated with reduced protection and the degree to which protection associated with seropositivity may actually be mediated by antibodies versus other forms (e.g. T-cell based) immunity.^6^

## Data Availability

The data is not publicly available.

## Acknowledgements

This work was supported by the National Cancer Institute, Office of the Director. We would like to thank Dr. Tony Fauci and Dr. Cliff Lane for comments on the manuscript. Authors RAH, JAR, CAK, WT are employees of Aetion. Authors SL, RK are employees of HealthVerity. Authors HK, WM are employees of Quest Diagnostics. And authors SA and OC are employees of LabCorp.

Funded by the National Cancer Institute

## Supplemental Materials. HealthVerity Data

HealthVerity data contain near real-time medical claims and outpatient pharmacy transactions, including drugs, diagnoses, procedures and selected lab results (incl. COVID testing). These data are drawn from a variety of US sources which include Veradigm and over 70 other HealthVerity data partners. Data elements include provider-submitted claims, adjudicated insurance claims, pharmacy billing managers claims submissions, and US laboratory chain test orders and selected results. They update in near real-time, with minimal lag between time of claim submission and time of inclusion in the database. 12+ months of historical data is available for many patients. Hospitalizations are included in the data at a summary level. Further, near real-time hospital chargemaster data, consisting of data from internal hospital billing systems, and capturing all billable drugs, procedures, and medical services provided to a patient in an in-hospital setting is available (without history) for select patients. Data are augmented for select patients with detailed clinical data from electronic health records. As information about co-morbidities and other risk factors is drawn from these data sources, recorded values may under-represent conditions that are not well-reported in insurance claims or require detailed historical hospital information. For reported baseline use of medications, drugs dispensed by a pharmacy are generally very well captured, though OTC medication use may not be

All data include key factors such as patient age, gender, and 3-digit zip level and may include an identifier for the treating provider. Data are de-identified and were certified HIPAA compliant by expert determination.

Death information is generally available with minimal lag for patients who die in a hospital setting, and for other settings, may also be linked from other data sources (with additional lag time).

Each dataset is provided by linking records on a unique patient identifier created by HealthVerity. The linkage of patients has high accuracy: 99.7% of linkage made are made correctly (0.3% false positives), and 96% of possible linkages are made (4% false negative). All linkage is done according to HIPAA regulations. With real-time assembly of data requiring the use of multiple sources, this approach appropriately balances timeliness with fidelity of linkage.

**Supplemental Materials Figure S1.**
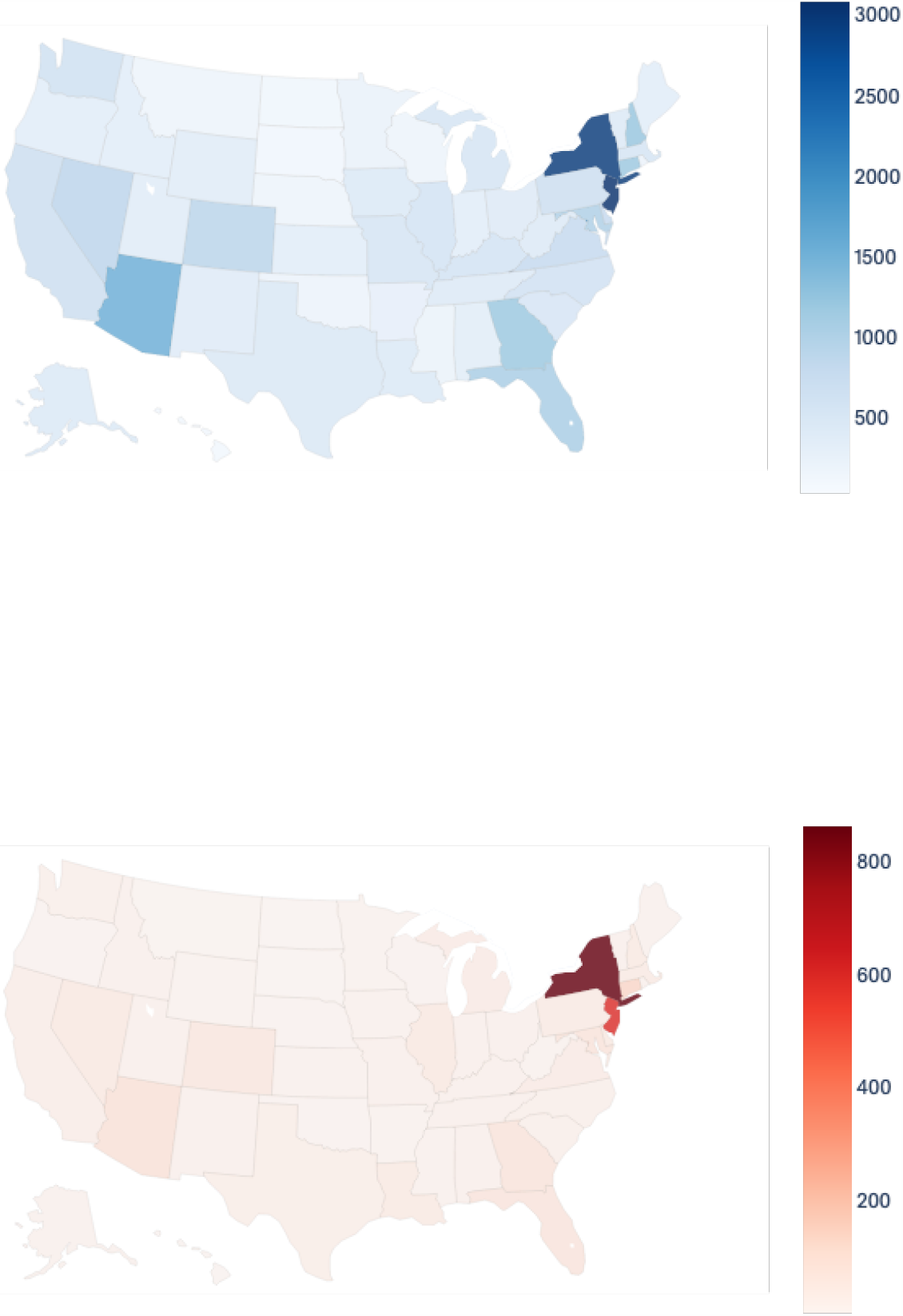
Rates of index seropositivity and seronegativity by US state. This figure shows the frequency of index seropositivity (red) and index seronegativity (blue) per 100,000 residents of each US state.

**Supplemental Materials Table S1.** This table provides a preliminary analysis of the ratio of NAAT positivity in index antibody positive versus index antibody negative patients by geography and time interval. Results are consistent with overall analysis.

| **Ratio of NAAT Positive Among A Index Antibody Positive and Negative By Geographic Region and Observation Period** |  |  |  |  |
| --- | --- | --- | --- | --- |
| **Region** | *Follow-up Period* |  |  |  |
|  | 0-30 days | 31 -60 days | 61-90 days | 90+ days |
| **Overall** | 2.85<br>(2.73 - 2.97) | 0.67<br>(0.6 - 0.74) | 0.29<br>(0.24 - 0.35) | 0.1<br>(0.05 - 0.19) |
| **Northeast** | 5.39<br>(5.01 - 5.80) | 1.86<br>(1.60 - 2.15) | 0.75<br>(0.58 - 0.98) | 0.22<br>(0.09 - 0.49) |
| **Midwest** | 3.89<br>(3.15 - 4.79) | 1.13<br>(0.67 - 1.92) | 0.63<br>(0.24 - 1.70) | 0<br>(0.0 - 0.0) |
| **South** | 2.14<br>(1.98 - 2.31) | 0.53<br>(0.39 - 0.72) | 0.36<br>(0.22 - 0.58) | 0.21<br>(0.05 - 0.84) |
| **West** | 3.62<br>(3.23 - 4.07) | 0.69<br>(0.44 - 1.07) | 0.46<br>(0.21 - 1.02) | 0<br>(0.0 - 0.0) |
| **Unknown** | 3.4<br>(2.93 - 3.95) | 0.62<br>(0.38 - 1.01) | 0.26<br>(0.11 - 0.58) | 0.31<br>(0.07 - 1.27) |

## References

1. Stadlbauer D, Tan J, Jiang K, et al. Repeated cross-sectional sero-monitoring of SARS-CoV-2 in New York City. Nature. Published online 2020:1–7.

2. Wajnberg A, Amanat F, Firpo A, et al. Robust neutralizing antibodies to SARS-CoV-2 infection persist for months. Science. Published online October 28, 2020:eabd7728. doi:10.1126/science.abd7728

3. Gudbjartsson DF, Norddahl GL, Melsted P, et al. Humoral immune response to SARS-CoV-2 in Iceland. N Engl J Med. 2020;383(18):1724–1734.

4. Long Q-X, Tang X-J, Shi Q-L, et al. Clinical and immunological assessment of asymptomatic SARS-CoV-2 infections. Nat Med. 2020;26(8):1200–1204. doi:10.1038/s41591-020-0965-6

5. Ward H, Atchison CJ, Whitaker M, et al. Antibody prevalence for SARS-CoV-2 in England following first peak of the pandemic: REACT2 study in 100,000 adults. medRxiv. Published online January 1, 2020:2020.08.12.20173690. doi:10.1101/2020.08.12.20173690

6. Zuo J, Dowell A, Pearce H, et al. Robust SARS-CoV-2-specific T-cell immunity is maintained at 6 months following primary infection. bioRxiv. Published online January 1, 2020:2020.11.01.362319. doi:10.1101/2020.11.01.362319

7. Ibarrondo FJ, Fulcher JA, Goodman-Meza D, et al. Rapid Decay of Anti–SARS-CoV-2 Antibodies in Persons with Mild Covid-19. N Engl J Med. 2020;383(11):1085–1087. doi:10.1056/NEJMc2025179

8. Pray IW. COVID-19 Outbreak at an Overnight Summer School Retreat―Wisconsin, July– August 2020. MMWR Morb Mortal Wkly Rep. 2020;69.

9. Addetia A, Crawford KH, Dingens A, et al. Neutralizing antibodies correlate with protection from SARS-CoV-2 in humans during a fishery vessel outbreak with high attack rate. medRxiv. Published online January 1, 2020:2020.08.13.20173161. doi:10.1101/2020.08.13.20173161

10. First Case of COVID-19 Reinfection Detected in the US. AJMC. Accessed November 11, 2020. https://www.ajmc.com/view/first-case-of-covid-19-reinfection-detected-in-the-us

11. Exchange JVLBVWN. Albany County resident reinfected with COVID-19 months after recovery. Casper Star-Tribune Online. Accessed November 11, 2020. https://trib.com/news/state-and-regional/health/albany-county-resident-reinfected-with-covid-19-months-after-recovery/article_b88aeff7-091e-5afb-9ee3-1b5715e9e219.html

12. Tillett RL, Sevinsky JR, Hartley PD, et al. Genomic evidence for reinfection with SARS-CoV-2: a case study. Lancet Infect Dis. Published online 2020.

13. Iwasaki A. What reinfections mean for COVID-19. Lancet Infect Dis. 2020;0(0). doi:10.1016/S1473-3099(20)30783-0

14. Hoang VT, Dao TL, Gautret P. Recurrence of positive SARS-CoV-2 in patients recovered from COVID-19. J Med Virol. Published online 2020.

15. Li N, Wang X, Lv T. Prolonged SARS-CoV-2 RNA shedding: Not a rare phenomenon. J Med Virol. Published online 2020.

16. AlJishi JM, Al-Tawfiq JA. Intermittent viral shedding in respiratory samples of patients with SARS-CoV-2: observational analysis with infection control implications. J Hosp Infect. Published online September 10, 2020:S0195-6701(20)30426-6. doi:10.1016/j.jhin.2020.09.011

17. Lee PH, Tay WC, Sutjipto S, et al. Associations of viral ribonucleic acid (RNA) shedding patterns with clinical illness and immune responses in Severe Acute Respiratory Syndrome Coronavirus 2 (SARS-CoV-2) infection. Clin Transl Immunol. 2020;9(7):e1160.

18. Li T, Cao Z, Chen Y, et al. Duration of SARS-CoV-2 RNA shedding and factors associated with prolonged viral shedding in patients with COVID-19. J Med Virol. Published online 2020.

19. Morone G, Palomba A, Iosa M, et al. Incidence and Persistence of Viral Shedding in COVID-19 Post-acute Patients With Negativized Pharyngeal Swab: A Systematic Review. Front Med. 2020;7.

20. Surkova E, Nikolayevskyy V, Drobniewski F. False-positive COVID-19 results: hidden problems and costs. Lancet Respir Med. Published online 2020.

21. Agarwal V, Venkatakrishnan AJ, Puranik A, et al. Long-term SARS-CoV-2 RNA shedding and its temporal association to IgG seropositivity. Cell Death Discov. 2020;6(1):138. doi:10.1038/s41420-020-00375-y

22. Meschi S, Colavita F, Bordi L, et al. Performance evaluation of Abbott ARCHITECT SARS-CoV-2 IgG immunoassay in comparison with indirect immunofluorescence and virus microneutralization test. J Clin Virol. 2020;129:104539. doi:10.1016/j.jcv.2020.104539

23. Health C for D and R. EUA Authorized Serology Test Performance. FDA. Published online December 7, 2020. Accessed December 7, 2020. https://www.fda.gov/medical-devices/coronavirus-disease-2019-covid-19-emergency-use-authorizations-medical-devices/eua-authorized-serology-test-performance

24. Sethuraman N, Jeremiah SS, Ryo A. Interpreting diagnostic tests for SARS-CoV-2. Jama. Published online 2020.

25. Ainsworth M, Andersson M, Auckland K, et al. Performance characteristics of five immunoassays for SARS-CoV-2: a head-to-head benchmark comparison. Lancet Infect Dis. 2020;20(12):1390–1400.

26. Theel ES, Harring J, Hilgart H, Granger D. Performance Characteristics of Four High-Throughput Immunoassays for Detection of IgG Antibodies against SARS-CoV-2. J Clin Microbiol. Published online 2020.

27. Hopkins JS. Pfizer’s Covid-19 Vaccine Proves 90% Effective in Latest Trials. Wall Street Journal. https://www.wsj.com/articles/covid-19-vaccine-from-pfizer-and-biontech-works-better-than-expected-11604922300. xPublished November 9, 2020. Accessed November 9, 2020.

28. Promising Interim Results from Clinical Trial of NIH-Moderna COVID-19 Vaccine. National Institutes of Health (NIH). Published November 15, 2020. Accessed December 11, 2020. https://www.nih.gov/news-events/news-releases/promising-interim-results-clinical-trial-nih-moderna-covid-19-vaccine

29. Polack FP, Thomas SJ, Kitchin N, et al. Safety and Efficacy of the BNT162b2 mRNA Covid-19 Vaccine. N Engl J Med. Published online December 10, 2020. doi:10.1056/NEJMoa2034577

30. Pottegård A, Kurz X, Moore N, Christiansen CF, Klungel O. Considerations for pharmacoepidemiological analyses in the SARS-CoV-2 pandemic#. Pharmacoepidemiol Drug Saf. Published online 2020.

31. Camm AJ, Fox KA. Strengths and weaknesses of ‘real-world’studies involving non-vitamin K antagonist oral anticoagulants. Open Heart. 2018;5(1).

32. Makady A, de Boer A, Hillege H, Klungel O, Goettsch W. What is real-world data? A review of definitions based on literature and stakeholder interviews. Value Health. 2017;20(7):858–865.

33. Miksad RA, Abernethy AP. Harnessing the power of real-world evidence (RWE): a checklist to ensure regulatory-grade data quality. Clin Pharmacol Ther. 2018;103(2):202–205.

